# Maternal Autoantibody Combinations as Predictors of Autism Risk: Retrospective Discovery and External Validation in a Prospective Cohort at Increased Risk

**DOI:** 10.64898/2026.10.23.26366364

**Authors:** E. Robert Wassman, Megan Martin, Joseph Schauer, Judy Van de Water, Michael Paul

**Affiliations:** Marabio Systems, Inc. (“MARAbio”), Salt Lake City, UT, 84101; University of California Davis, Department of Internal Medicine and UC Davis MIND Institute, Sacramento, CA, 95817

**Keywords:** Autism, ASD, Maternal Autoantibody Related Autism (MARA), indirect ELISA, LDH-A, LDH-B, GAH, STI1, CRMP-1, CRMP-2, NSE and YB-1, CHARGE, MARBLES

## Abstract

**Background/Objectives:** Maternal Autoantibody-Related Autism (MARA) is a recently defined subset of autism characterized by specific combinations of maternal autoantibodies which cross the placenta and bind neurodevelopmentally critical target proteins. A clinical multi-ELISA MARA assay was recently rigorously analytically validated. The primary objective of this study was to assess the potential performance of this MARA autoantibody assay for predicting autism risk in high-risk populations. Secondary objectives were to establish the significant MARA autoantibody combinations and verify clinical discrimination in an external cohort.

**Methods:** Bio-banked plasma samples from mothers of 420 autism and 335 typically developing (TD) children were analyzed by ELISA for MARA autoantibodies in combination. “Transcendent” autoantibody pairwise combinations were identified in discovery where both autoantibodies in a combination exceeded the values of every control participant. Permutation testing was used to establish that the probability of observed results significantly exceeded chance. Performance was then validated in an independent cohort (mothers of 42 autistic and 90 TD children).

**Results:** “Transcendent” antibody levels were discovered in 70 mothers (16.7% of autism cases; no typically developing participants) in the discovery cohort. Specificity and positive predictive values for autism were 100% by construct. An external cohort showed similar performance with 14 (33.3%) transcendents. Permutation testing of each cohort demonstrated probabilities of transcendent cases to be significantly higher than expected be chance.

**Conclusions:** The data reported here provides evidence for a high-risk subset of mothers of children with autism identified by the MARA autoantibody assay which enables further clinical validation.

## Introduction

Autism spectrum disorder (Autism) is a childhood-onset neurodevelopmental disorder of variable severity characterized by difficulties with social interaction and communication and restricted or repetitive patterns of thought and behavior. The prevalence in US children has risen dramatically over the past two decades, from 1 in 150 in 2000 to 1 in 31 in 2022, making autism an important health concern and a substantial socioeconomic burden for affected families and the healthcare system [1,2]. The development of autism is strongly influenced by genetics and environmental factors, including the maternal immune system during gestation [3–5].

Although clinical diagnosis remains the gold standard for autism and can be highly accurate, it requires comprehensive clinical and psychological assessment, and children typically do not receive a definitive diagnosis until approximately 3–5 years of age [6]. Consequently, this prolonged diagnostic path often delays access to early intervention and support [6]. The identification and validation of biomarkers that are mechanistically linked to the underlying biology of autism can provide a critical clinical tool for early diagnosis.

Maternal autoantibodies targeting neurodevelopmentally relevant fetal brain proteins were first described in 2008 by investigators in the Van de Water laboratory at the Medical Investigation of Neurodevelopmental Disorders (M.I.N.D.) Institute at the University of California, Davis [7]. Maternal IgG antibodies routinely traverse the placenta and, during critical periods of fetal brain development, specific autoantibodies may gain access to antigens within the developing brain [8,9]. This possibility is particularly relevant during stages of gestation when the blood–brain barrier is relatively permissive, providing a potential mechanism through which maternal autoantibodies may influence neurodevelopment [8,9].

Evidence from animal models provides support for a potential pathogenic role of MARA autoantibodies beyond their association as biomarkers for autism. Studies using both passive antibody transfer and active induction of maternal autoantibody production have demonstrated autism-relevant behavioral phenotypes and neuroanatomical alterations in offspring following gestational exposure to these autoantibodies [10–16]. These findings raise the possibility that MARA autoantibodies represent mechanistic biomarkers rather than being solely correlational markers of autism risk.

Epidemiologic studies have demonstrated that the presence of maternal autoantibodies directed against specific combinations of eight proteins that are highly expressed during brain development are strongly associated with an increased likelihood of autism in offspring [5,7,17–22]. Maternal Autoantibody-Related Autism (MARA) patterns have been estimated to occur in 10 to 24% of mothers associated with autism [17,22]. The Childhood Autism Risks from the Genetics and Environment (CHARGE) study was a large case–control investigation of underlying genetic and environmental causes for autism [23]. In this study, MARA autism-specific patterns demonstrated greater than 99% classification accuracy and identified potential risk biomarkers associated with a subset of up to 18% of autism cases [7,17,24]. An independent clinical validation using the prospective Early Markers for Autism (EMA) cohort closely replicated these findings and importantly demonstrated that the potentially pathogenic autoantibodies are present during pregnancy, before the birth of the affected child [21].

Several recent comprehensive reviews of MARA also address maternal autoantibody correlation to neurodevelopmental disorders. A book chapter by Van de Water and Wassman reviewed many preclinical and human studies conducted in academic laboratories, providing extensive evidence that maternal autoantibodies reactive to specific fetal brain proteins may be highly specific biomarkers of autism risk [25]. Also an independent comprehensive review of maternal autoantibodies, which extensively evaluated the work of Van de Water and colleagues defining MARA as well as other, less extensively characterized autoantibodies, concluded that maternal autoantibody status is associated with autism risk in offspring, and these autoantibodies are potentially modifiable environmental contributors [26].

Rigorously translating these mechanistic biomarkers into clinical application provides an important avenue toward earlier identification of autism and, potentially, the development of targeted therapeutic or preventive interventions for a subset of autistic individuals, i.e., personalized immunology. Recently we reported the analytical validation of enzyme-linked immunosorbent assays (ELISAs) to rigorous standards to establish assay performance suitable for their intended clinical use in detecting MARA autoantibodies [27]. The antigens used in these ELISAs were Collapsin response mediator proteins 1 and 2 (CRMP-1 and CRMP-2), Guanine deaminase (GAH), Stress-induced phosphoprotein 1 (STI1), Neuron-specific enolase (NSE), L-lactate dehydrogenase A and B chains (LDH-A and LDH-B), and Y-box-binding protein 1 (YB-1), all of which had been identified as MARA autoantibody antigens in the academic studies described in the literature [5,7,17–22,27]. These assays were then validated independently in two Clinical Laboratories Improvement Act (CLIA) and College of American Pathologists (CAP) certified laboratories to clinical laboratory standards [27].

The primary objective of this study was to demonstrate the ability of the MARA autoantibody assay for predicting the risk of autism for intended uses (IU) in two high-risk populations: 1) women with a child who manifests specific signs/symptoms of autism (IU1), and 2) women with a child previously diagnosed with autism prior to a subsequent pregnancy, or before a subsequent child develops signs/symptoms of autism (IU2), based on the discovery and evaluation of “transcendent” autoantibody levels in a subset of high-risk mothers where both autoantibodies in a combination exceed the values of every control participant. A secondary objective was to establish the statistically most significant MARA autoantibody pairwise combinations as the foundation for subsequent development and validation of an ELISA-based clinical assay test table.

The work presented here builds on the previous academic studies that used an early, less standardized prototype of the eight-autoantibody ELISA assay to demonstrate preliminary risk assessment [5,7,17–19,22]. MARA autoantibody assays which utilized fixed thresholds corresponding to the 97.5th percentile of a normal population distribution generated from blood bank samples, as well as previously identified pairwise autoantibody combinations similar to those reported in the academic literature [27]. However, this approach could not identify sufficiently robust thresholds to achieve clinically useful discrimination of autism from typical development. The current study was designed to repeat the classifier discovery process using an analytically validated assay performed in a clinical laboratory to evaluate all possible pairs of autoantibodies, and identify potential criteria which would yield a very low false positive rate, potentially suitable for transition to clinical application. The discovery of transcendents met these criteria and this study evaluated the characteristics of the transcendent population and its performance to discriminate a MARA subset of mothers of children with autism.

## Materials and Methods

### Study Design

This was a retrospective discovery and validation study using banked maternal plasma samples with supporting clinical data from the CHARGE cohort assayed in a CLIA/CAP-certified laboratory for the presence of eight autoantibodies: LDH-A, LDH-B, GAH, STI1, CRMP-1, CRMP-2, NSE and YB-1.

This analysis proceeded through multiple stages. In the discovery stage, an extensive analysis of the CHARGE data set (N =755) was deployed to identify the relationship between autoantibody assay results and clinical status of the mother’s children. At this point a clear set of mothers of children with autism was identified with “transcendent” autoantibody levels for specific pairwise combinations.

A mother was defined as transcendent for a given autoantibody combination when her ELISA assay values for both members of the combination exceeded the corresponding values of every control participant (mothers of TD children). Permutation testing with randomized case–control labels was used to establish, both for the overall group of transcendent cases and for each individual combination, whether observed ELISA counts exceeded the 95th percentile of counts arising by chance. In the discovery phase, a risk histogram for the CHARGE cohort was generated to evaluate the distribution of risk for autism and TD participants and to compare the accuracy of the maternal autoantibody pairwise combination assay as a predictor of autism in the offspring versus the previously determined gold standard clinical diagnosis (predicted risk versus actual risk). These were further assessed by contingency matrix, and the Fisher Exact Test.

Following the discovery phase, the autoantibody pairwise combinations identified in the discovery phase were externally validated in a small external cohort from the Markers of Autism Risk in Babies—Learning Early Signs (MARBLES) cohort, which consisted of samples from mothers of previously diagnosed children with autism having another child (42 subsequent children with autism, and 90 TD children). Similarly, using the same approach as with the discovery cohort, a set of transcendent participants was identified and similarly assessed in this independent population.

### Specimen Sources

In the discovery phase, bio-banked plasma samples sourced from the CHARGE study were used [23]. These were collected in acid–citrate–dextrose (ACD) tubes (BD Diagnostics) from mothers of 420 autism and 335 typically developing (TD) children. In the external validation stage of the study, 132 bio-banked maternal ACD plasma samples from the MARBLES study (42 subsequent offspring with autism, 90 with TD) [28] were assessed. Participants were originally selected based on the inclusion/exclusion criteria of the CHARGE and MARBLES studies, and samples were collected, processed and stored as described in the respective protocols [23,28].

Only bio-banked patient samples were used in this study. Both the previously conducted CHARGE and MARBLES studies complied with all applicable regulatory requirements regarding human participants, including informed consent obtained before collection of any data, and approval by the institutional review boards for the State of California and the University of California [23,28].

CHARGE was a time perspective case-control study designed to assess genetic and environmental factors contributing to autism, which enrolled children aged 24 to 60 months and their mothers between 2003 and 2011, recruited through the California Department of Developmental Services and the Medical Investigation of Neurodevelopmental Disorders (MIND) Institute at the University of California, Davis. [23]. The subject inclusion criteria for the CHARGE participant subset was mothers of children diagnosed with autism by gold standard clinical diagnostic protocols at the MIND Institute/UC Davis. TD controls were randomly selected from birth records and frequency matched on age, sex, and broad geographic region. Children recruited from the general population were confirmed as TD based on the Social Communication Questionnaire [29]. The CHARGE cohort originally collected specimens from mothers of 450 autism and 343 TD children and all individuals where sufficient volume remained in the MIND biobank were included in this study with no additional selection other than mothers of children without autism.

The study population for MARBLES was mothers who already had a child or children diagnosed with autism by gold standard clinical criteria and who were pregnant or planning a pregnancy [28]. Recruitment was primarily through the California Department of Developmental Services or the UC Davis MIND Institute. Estimates indicate that about 75–80% of affected children in California are enrolled in these services, however milder cases would be underrepresented because of the eligibility requirements for DDS participation [28,30]. Selection criteria of MARBLES samples for this study were those with maternal samples at or beyond 24-months post-delivery selected from all those where a gold standard clinical autism evaluation and an intellectual disability (ID) evaluation (irrespective of either outcome) had been completed. The 132 available samples used in this study represented outcomes of autism in 42 of the subsequent offspring and TD in the remaining 90 offspring.

Children with other developmental concerns in both cohorts were excluded from this analysis. Also, MARBLES samples collected during pregnancy were excluded for this analysis as the MARA autoantibody assay is not yet analytically validated for use on maternal samples during pregnancy. Twenty women with affected children diagnosed during the CHARGE study also participated in the MARBLES study based on the diagnoses of autism in the initial child, however none of these were selected for inclusion based on their prior (CHARGE) MARA test results. These cases were excluded from analysis in the CHARGE discovery cohort for temporal blinding to ensure there was no overlap between the discovery and validation set, and thus only utilized in the MARBLES external validation analysis.

Clinical diagnosis of autism at the MIND Institute with gold standard criteria served as the reference standard for both cohorts for evaluating test accuracy. Demographic data and diagnoses collected within the CHARGE and MARBLES studies were used during the analysis of the results of this study.

### Blinding

De-identified study samples were assayed blindly at the CLIA laboratory. Permutation analyses were unblinded as these processes obviate the need for blinding. MARBLES samples remained blinded until final external validation study completed. MARAbio Clinical, Development, and Research personnel were blinded to the study results until the analysis was completed.

### MARA Autoantibody ELISA Procedure Methodology

All maternal blood samples had been collected in 8 ml ACD tubes (BD Diagnostics) and plasma was separated, labeled, aliquoted, and stored at −80 °C within 24 hours of drawing. Banked samples from CHARGE and MARBLES that had been frozen and thawed more than twice were excluded.

The ELISA procedure methodology was performed as described previously in detail [27]. Briefly, all assays were indirect ELISAs with recombinant protein antigens (LDH-A, LDH-B, GAH, ST1, CRMP-1, CRMP-2, NSE, or YB-1) (sourced from Expression Systems, LLC, Davis, CA, USA) bound to microarray plates, incubated with the test maternal plasma samples, Goat anti-human IgG horseradish peroxidase (HRP)-conjugated secondary antibody, and finally a chromogenic substrate prior to reading optical density (OD) on a spectrophotometer at 450 nm with a reference at 595 nm. A single point calibration method was used along with a reagent blank control. In positive samples, blue color developed in the wells at an intensity proportional to the concentration of protein-specific antibodies.

### Definition of Transcendent Values

The ELISA assay result of a mother of a child with autism was defined as *transcendent* if, for any given autoantibody combination the values for both autoantibodies in the combination exceeded the corresponding values of every control participant in the cohort. Transcendence is therefore a property of a participant’s sample with respect to a pairwise combination, defined entirely by observed complete separation from the control distribution, i.e., equivalent to setting a classifier decision threshold at the maximum value of the true-negative samples in the training dataset, which may reduce reliance on a fitted threshold, or optimization against a chosen operating point, and no free parameters. A participant’s sample is counted once as a transcendent case if it is transcendent for at least one pairwise combination without regard here for situations where multiple combinations were present in the same individual. Because the criterion requires that no control participant exceed the threshold for either antigen, specificity within the cohort in which transcendence is assessed is 100% by construction, and positive predictive value is correspondingly 100%. The informative quantities are therefore the number of transcendent cases observed, the distribution of those cases across pairwise combinations, and whether either exceeds what would be expected by chance.

For both the CHARGE and MARBLES cohorts, for each of the 28 pairwise autoantibody combinations, every autism participant was evaluated against the transcendence criterion defined above and the total number of transcendent autism cases was determined. Three figures of merit were specified in advance: the total transcendent cases in the population tested; the distribution of those cases across pairwise combinations, displayed as a heatmap; and the resulting sensitivity, specificity, PPV, and NPV that were computed over the whole cohort. We note that by definition of transcendence, there are no false positives, and so, specificity and PPV will be 100% by definition. Statistical significance was assessed by permutation testing, which establishes whether the observed number of transcendent cases is higher than would be expected by chance.

### Permutation Analysis

We performed permutation testing to test that the model performance metrics was significantly better than chance. A non-parametric test, permutation testing empirically defines the null distribution by randomly shuffling the data labels, so does not require any distributional assumptions and provides a powerful and interpretable assessment of significance even when other methods may be inappropriate. The data labels were randomly shuffled within each cohort (MARBLES or CHARGE) 10,000 times while preserving the case-control ratio. For each iteration the transcendence criterion was applied to the shuffled data, and the resulting number of transcendent cases was recorded both overall and for each individual autoantibody combination[31]. Autoantibody combinations with significant signal show actual observed counts of transcendent cases greater than the 95th percentile of counts found in the data with shuffled labels. Additionally, note that the labels are shuffled separately within CHARGE and within MARBLES so that any effects present due to differences in the characteristics of samples from the two cohorts are preserved through the permutation. To assess the utility of a subpanel of the 28 pairwise combinations, the optimal panel of five combinations, in terms of identification of transcendents across CHARGE and MARBLES, was derived. The number of transcendents identified by this optimal panel were determined and statistical significance determined by permutation testing.

### External Validation on the MARBLES Cohort

An external validation was subsequently performed on the MARBLES cohort (N =132; 42 mothers whose subsequent child was diagnosed with autism and 90 whose subsequent child was TD). All MARBLES mothers who had bio-banked samples around the 24-month post-partum timepoint were assayed in the same CLIA/CAP-certified laboratory under the same analytical protocols. The external validation cohort was independent of the CHARGE cohort, with the exception of the twenty women with affected children diagnosed during the CHARGE study who enrolled in MARBLES based on those diagnoses but in this study were only considered in their MARBLES context. Verification of transcendence proceeded in the same manner as discovery on CHARGE by generation of the same three figures of merit: the total transcendent cases, the resulting performance metrics, and permutation testing of significance. Permutation testing was performed to assess whether the observed model performance significantly exceeded that achievable by chance alone. The Fisher Test of significance was used to assess the significance of the number of transcendents discovered in the context of discrimination of autism and TD participants. Validation was specified in advance as successful if transcendent cases were again observed in MARBLES and if their number exceeded the 95th percentile of the corresponding permutation distribution.

### Risk Score Distributions

Risk score distribution was used here to compare the accuracy of the assay for maternal autoantibody pairwise combinations as a predictor of autism in the mothers’ offspring to the gold standard clinical diagnostic methodology for CHARGE participants and separately later for the MARBLES cohort. Predicted risk is the maximum PPV derived from that participant’s assay values for any pairwise combination. Actual risk is the rate of autism cases based on gold standard clinical diagnosis.

To understand how pairwise combinations of autoantibodies stratify participants in terms of autism risk, we defined a risk score “Risk(s)” for every participant. To determine this, for a combination of autoantibodies P1 and P2, we used the MARA-autoantibody assay values for participants, and T1 and T2, as decision thresholds. Test positive subjects were defined as the proportion of participants with assay values that are equivalent to or exceed that of T1 and T2,and the remainder were designated as test negative. Next we defined “Risk(s)” to be the maximum risk observed across all 28 autoantibody pairwise combinations. Note that a participant has a Risk score of 100% if and only if it is transcendent.

This definition of risk allows us to plot the distribution of all participants in each cohort by risk score as derived from the MARA-autism assay, and importantly, plot the distribution of risk scores for TD versus autism participants as a normalized grouped histogram.

### Statistical Analysis of Performance

Sensitivity was calculated as the number of transcendent autism cases divided by the total number of autism participants in the cohort, and specificity as the proportion of control participants not meeting the transcendence criterion. Positive and negative predictive values were computed from the prevalence of the cohort from which they were collected and are not transferable to a population of different prevalence. 95% CIs for sensitivity and transcendent counts were computed with the exact Clopper–Pearson method [32]. Because predictive values in a case-control design depend on the sampled case fraction, NPV CIs were computed by the logit method of Mercaldo et al.[33], with prevalence fixed at each cohort’s observed case proportion. As defined, transcendence requires complete separation from the control distribution, therefore specificity and PPV equal 100% by construction, therefore no confidence interval is reported for them; the inferential weight is carried instead by the permutation comparison described above.

For calculations of PPV and NPV, the observed prevalence reported for the two study populations were ∼55% for CHARGE and 32% for MARBLES.

The Fisher exact test was used to test for significance of association for both CHARGE and MARBLES cohorts.

## Results

### Identification of a Transcendent Risk Population from CHARGE

Across all 28 pairwise autoantibody combinations, 70 transcendent autism cases (95% CI 56-86) were identified in the CHARGE cohort, corresponding to a sensitivity of 16.7% (70/420, 95% CI 13.2-20.6). Because transcendence requires that a case exceed every control participant on both antigens of a pairwise combination, no control participant met the criterion yielding both specificity and positive predictive value of 100% based on this definition rather than from calculation. The negative predictive value was 48.9% (95% CI 47.8-50.0). Permutation testing demonstrated that 70 transcendent cases is significantly higher than expected by chance, as described in “Permutation Testing” section below. The distribution of transcendent cases across pairwise combinations is shown in **Figure 1**.

**Figure 1.**
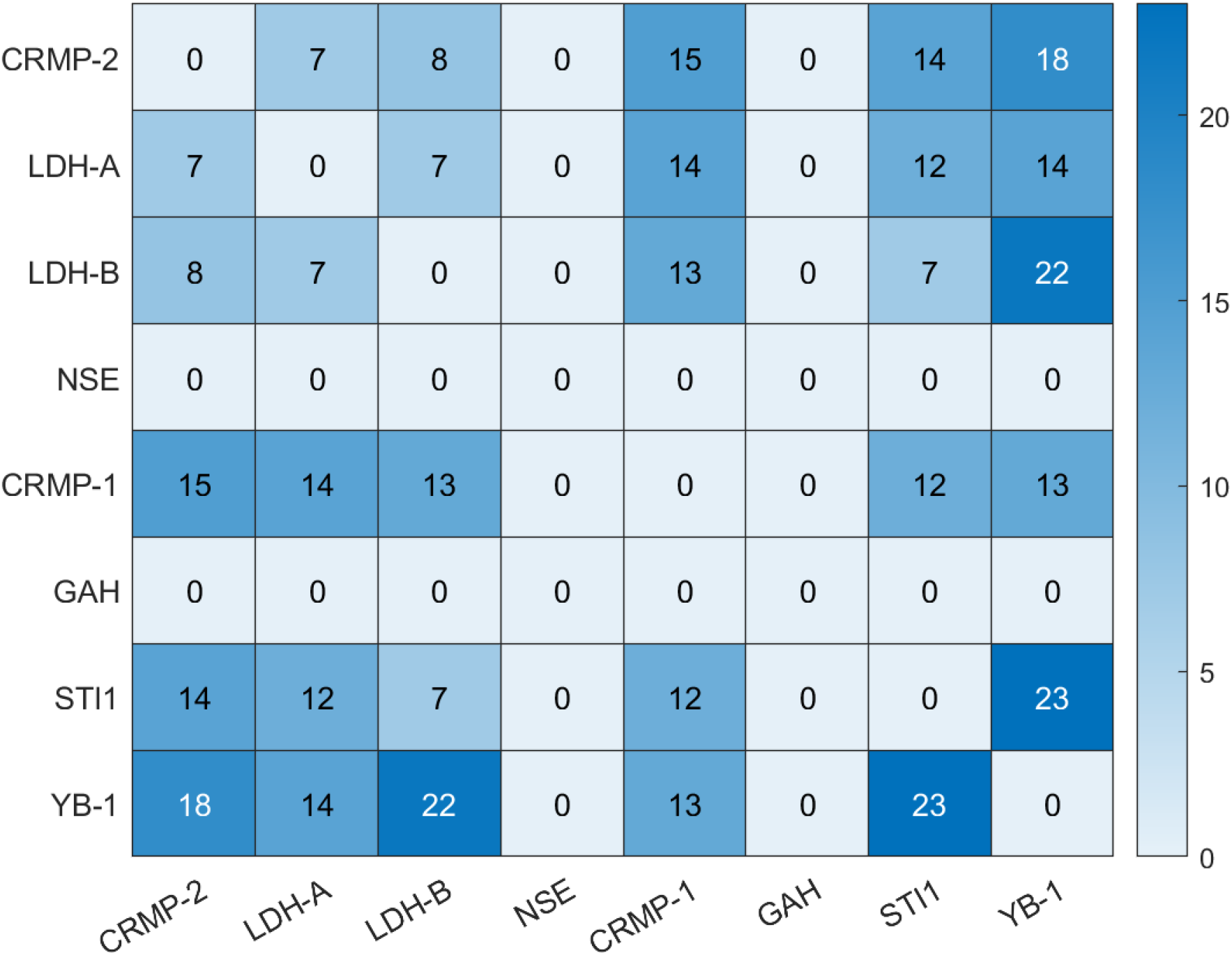
Transcendent autism cases per autoantibody combination in the CHARGE discovery cohort. Each cell displays the number of mothers of autistic offspring that are transcendent for the corresponding pair of autoantibodies (ELISA assay values for both antigens exceed those of every control participant)

### Contingency Matrix Derived from CHARGE Transcendent Observations

Across all pairwise combinations, from those demonstrably associated with transcendence in the CHARGE cohort, a contingency matrix was constructed **(Table 1).** All transcendence-positive participants had an autism diagnosis in their offspring and none of the TD participants were transcendent for MARA autoantibodies. **(Table 3)**. The Fisher Exact test applied to the contingency table yields a p < 0.001, signifying a statistically significant association between the variables.

**Table 1:** CHARGE Contingency Matrix. Resulting calculated performance in the CHARGE cohort using the transcendent definition for a positive test result were sensitivity: 16.7% (95% CI 13.2-20.6); specificity: 100%; PPV: 100%; and NPV: 48.9% (95% CI 47.8–50.0) (Table 3).

|  | AUTISM | TD |
| --- | --- | --- |
| Transcendent + | 70 | 0 |
| Transcendent - | 350 | 335 |

### External Validation on MARBLES

In order to validate the performance of the assay in identifying transcendents in an independent cohort, the MARBLES cohort was evaluated across all combinations of tested autoantibodies. The MARBLES cohort is distinct from the CHARGE cohort used in training in that it assesses maternal autoantibodies in relation to the subsequent clinical diagnosis of autism in a prospectively ascertained younger sibling of a previously diagnosed autistic child born to the same biological mother. Twenty mothers who participated in both studies were only analyzed in the context of MARA in the MARBLES cohort. Overall, individual autoantibody assay results across the eight autoantibodies and all combinations were highly consistent with CHARGE. Similar to CHARGE, a set of transcendent participants was identified in the independent MARBLES cohort. Fourteen transcendents (95% CI 8–21) were identified across all combinations, and all 14 were in mothers whose subsequent offspring went on to be diagnosed with autism. None of the resulting test-negative mothers had a subsequent child with an autism diagnosis. By permutation testing, the observed count of 14 transcendent cases fell in the 95th percentile of the null distribution demonstrating that the finding of these 14 transcendent cases is significantly higher than expected by chance. The Fisher Exact test applied to the corresponding contingency table **(Table 2)** yielded a p < 0.001, signifying a statistically significant association between the variables.

**Table 2:** MARBLES Contingency Matrix. Resulting calculated performance in the MARBLES cohort using the transcendent definition for a positive test result were sensitivity: 33.3% (95% CI 19.6-49.5); specificity: 100%; PPV: 100%; and NPV: 76.3% (95% CI 72.2–79.9) (Table 3).

|  | AUTISM | TD |
| --- | --- | --- |
| Transcendent + | 14 | 0 |
| Transcendent - | 28 | 90 |

### Permutation Testing

A test of significance (permutation testing) was done to confirm that the number of transcendent autism cases for both the CHARGE and MARBLES cohorts as well as the distribution of those cases across pairwise combinations exceeded what would arise by chance. Permutation testing applied 10,000 times to the same data randomized to assess how likely the pairwise combinations and high-risk results could be obtained by chance alone.

Permutation testing placed the 95th percentile of the null distribution at 51.5 transcendent cases, well below the 70 observed. The number of transcendent autism cases in CHARGE is therefore greater than would be expected by chance.

The optimal panel of five combinations, in terms of identifying the most transcendents across CHARGE and MARBLES was CRMP-1 + CRMP-2, LDH-A + STI1, CRMP-2 + YB-1, LDH-B + YB-1 and STI1 + YB-1. This panel identified 60 out of 70 transcendents in CHARGE and 9 of 14 transcendents in MARBLES, for a total of 69 transcendents out of a possible 84. By permutation testing, the 95^th^ percentile of transcendents identified by chance was 46. Consequently, this optimal panel is statistically significant for identification of transcendents.

### Risk Score Distributions

To compare the accuracy of the assay for maternal autoantibody combinations and our defined transcendent threshholds as predictors of autism risk in the mothers’ offspring to their respective gold standard clinical diagnoses, the distribution of predicted risks for participants in each cohort was plotted in normalized grouped histograms of TD and autism cases. **[Figures 2 (CHARGE) and 3 (MARBLES)]** The distribution of predicted risk across both cohorts is bimodal with a cluster at the far right of the graph where actual and predicted risk both equal 1.0. In CHARGE, this represents 70 extreme transcendent risk participants (16.7% of autism mothers), clearly separated from the remainder of cases and controls with no overlaps. This cluster observed at the high-risk end of the histogram was much higher than expected and met the definition of transcendents. The bulk of both CHARGE cases and controls fell between 0.55, i.e., approximately the cohort’s autism prevalence, and 0.95, while the isolated cluster of transcendents appears at predicted risk of 1.0, distinctly separated by a gap from the main distribution. Aside from this observed set of transcendents, which were all from mothers of children with autism, the distribution of cases and controls were not distinguishable.

**Figure 2.**
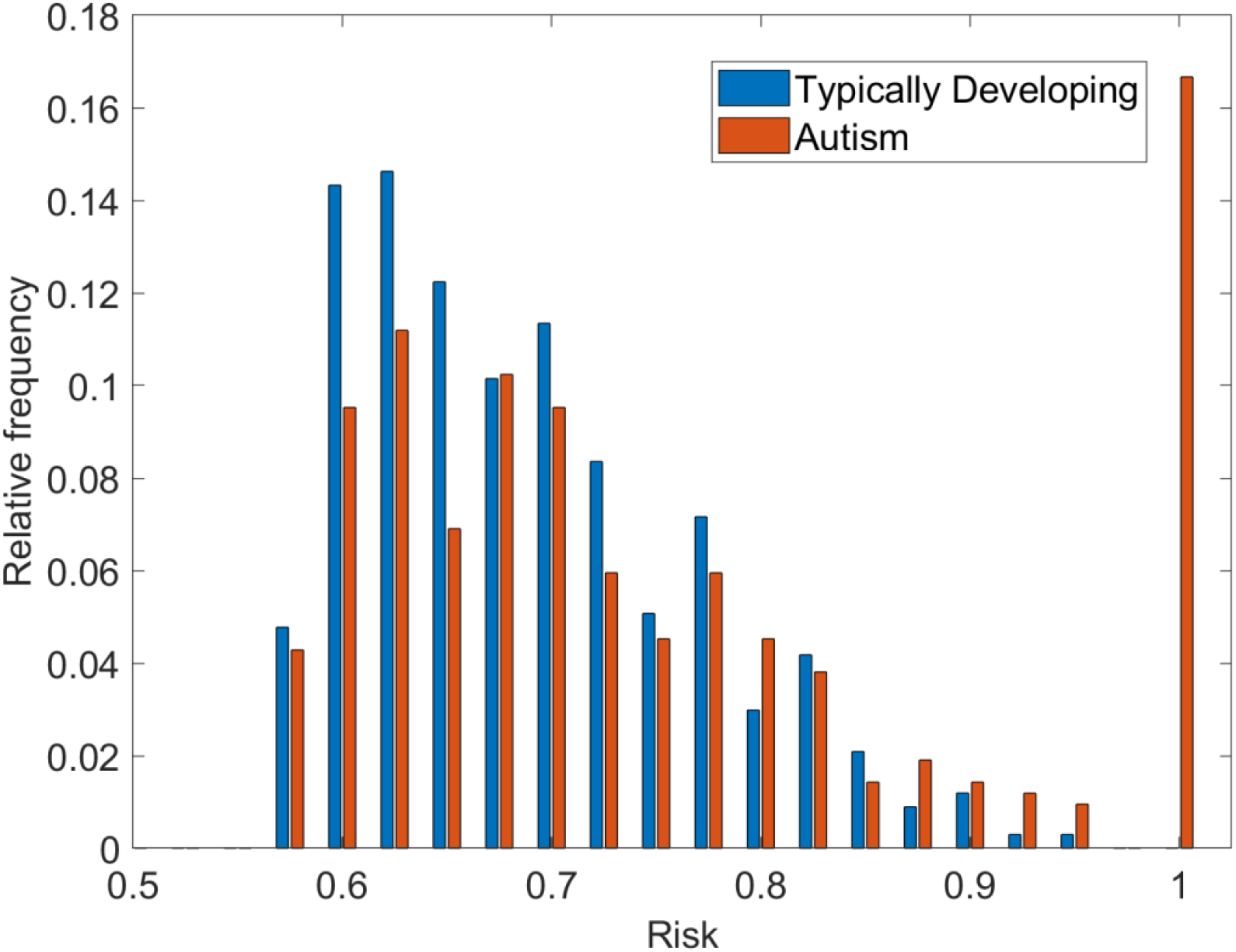
Distribution of predicted risks for the CHARGE participants, displayed in a normalized grouped histogram of TD and autism cases. The predicted risk of a participant was determined by taking their ELISA assay values for all 28 autoantibody pairwise combinations and using these as the thresholds and calculating the rate of autism at these thresholds. This highlights the higher than expected number of autism participants at risk level of 1, or 100%.

**Figure 3.**
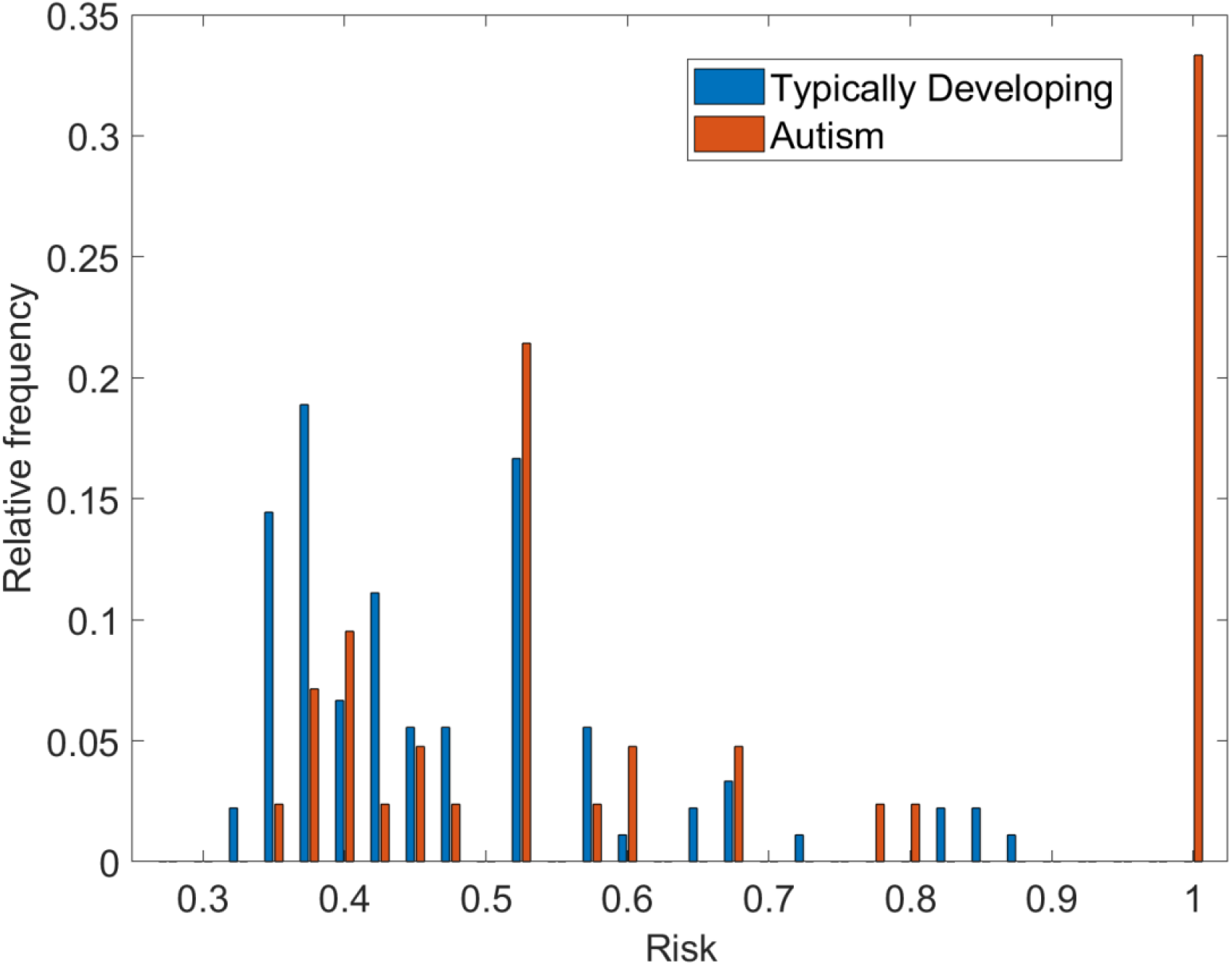
Distribution of predicted risks for the MARBLES participants, displayed in a normalized grouped histogram of TD and autism cases. The predicted risk of a participant was determined by taking the ELISA assay values for all 28 autoantibody pairwise combinations and using these as the thresholds and calculating the rate of autism at these thresholds. This highlights the higher than expected number of autism subjects at risk level of 1, or 100%.

In the MARBLES cohort, a similar gap exists between the 14 observed transcendents (33.3% of autism mothers) at a risk score of 1, however the range of risks for those below this was broader, ranging from 0.32 (again approximating that cohort’s autism prevalence) and 0.88. All of the transcendent results were from mothers whose subsequent child was diagnosed clinically with autism subsequently, while similarly to what was observed in CHARGE, no mothers with subsequent TD offspring had transcendent autoantibody levels. While the smaller numbers in this cohort make this histogram more subject to sampling variability, aside from this observed set of transcendents, the distribution of cases and controls were not distinguishable.

### Projected Performance Characteristics

The performance calculations from the CHARGE discovery cohort and the external validation in MARBLES are summarized in **Table 3**. In both cohorts, the presence of transcendent cases drives extremely high specificities and positive predictive values for the assay by definition. By permutation analysis, the observed counts of transcendents exceeded the 95% of the number expected by chance. The difference in detection rate, i.e., sensitivity in this context, between the two populations is likely partly a function of their size differences, but in MARBLES the denominator for sensitivity is children of mothers with a previously affected child versus mothers of all autistic children (in CHARGE). Regardless, both are within the previously observed metrics with these assays [17,18,21].

**Table 3.** Transcendence in discovery and external validation cohorts. Specificity and PPV are 100% by construction, since transcendence requires that no control participant exceeds the case on either antigen of a pairwise combination. Predictive values are reported based on the case-control composition of each cohort and are not translatable to populations with different prevalences. The optimal panel of five combinations identifying the most transcendents across CHARGE or MARBLES was CRMP-1 + CRMP-2, LDH-A + STI1, CRMP-2 + YB-1, LDH-B + YB-1 and STI1 + YB-1.

| **Metric** | **Discovery:**<br>**CHARGE (420 autism /**<br>**335 TD)** | **External validation:**<br>**MARBLES (42 autism /**<br>**90 TD)** |
| --- | --- | --- |
| Transcendent autism cases, N (95% CI) | 70 (56-86) | 14 (8-21) |
| Sensitivity, % (95% CI) | 16.7% (70/420)<br>(13.2-20.6) | 33.3% (14/42)<br>(19.6-49.5) |
| Specificity | 100% | 100% |
| PPV | 100% | 100% |
| NPV, % (95% CI)<sup>a</sup> | 48.9% (47.8-50.00) | 76.3% (72.2-79.9) |
| Transcendents captured by optimal five-pair panel | 60 transcendent cases | 9 transcendent cases |

## DISCUSSION

An immune-mediated subset of autism was previously identified in the Van de Water laboratory that was characterized by specific combinations of maternal autoantibodies which target proteins highly expressed in the developing brain [7]. A series of independent epidemiological studies as well as critical animal studies pointing to the pathogenicity of these antibodies further defined this subset known as maternal autoantibody related autism, or MARA [5,7–9,11–20,24,26]. These mechanistic biomarkers were recently translated into a clinical laboratory following optimization, standardization and analytical validation [27]. Clinical application of these biomarkers may enable earlier diagnosis and inform potential therapeutic or preventative strategies for this subset of autism, and the objective of the current study was to better determine their potential clinical predictive value, and set the foundation for developing and clinically validating a deployable test table in subsequent work.

The results from the large CHARGE cohort presented here closely recapitulated relevant pairwise autoantibody combinations and apparent predictive performance reported in multiple academic publications [17,18]. A key discovery finding was the presence of a substantial subset of mothers with autoantibody assay results which were defined as transcendent, i.e., for any given autoantibody combination the values for both autoantibodies in the combination exceeded the corresponding values of every control participant in the cohort. Transcendents by their definition maximize discrimination of mothers of children with autism from those with TD children and have specificities and PPVs of 100% by construct.

The findings in the CHARGE cohort were externally validated using a cohort (MARBLES) that was not involved in the discovery process, and in which maternal samples were obtained prospectively, before any diagnostic evaluation of a subsequent child. A similar percentage of transcendents were identified in this independent prospective cohort. Overall, there was significant agreement in the performance metrics between the CHARGE and MARBLES cohorts. These results provide additional evidence for clinical validity in both the CHARGE cohort, i.e., mothers of children diagnosed with autism, and the MARBLES populations, i.e. mothers with subsequent offspring following a confirmed diagnosis in a previously diagnosed sibling, and overall provide support for MARA as a distinct subset of autism.

In both the CHARGE discovery cohort and the MARBLES external validation cohort, all of the MARA positive mothers had children diagnosed subsequently with autism and none of the TD children’s mothers were MARA positive. Permutation testing and risk distribution analysis supported the assay’s significance in prediction of risk relative to the known gold standard clinical diagnoses. In our risk distribution analysis on the CHARGE cohort, the subset of 70 (16.7%) mothers of affected children with extreme (transcendent) risk values formed a distinct cluster, completely separated from the remainder of the control and autism populations, which otherwise exhibited fully overlapping population distributions. Similarly, the MARBLES cohort had a distinct cluster of 14 (33.3%) mothers of children with extreme (transcendent) risk values.

Five statistically significant combinations, CRMP-1 + CRMP-2, LDH-A + STI1, CRMP-2 + YB-1, LDH-B + YB-1 and STI1 + YB-1, contributed the most transcendents to the overall total number of transcendents in both cohorts (60/70 in CHARGE and 9/14 in MARBLES) for identifying the sub-population of mothers at very high risk for autism in their offspring, both current and future. Notably, three of the five, CRMP-1 + CRMP-2, LDH-B + YB-1, and STI1 + YB-1, are among the MAR autism-specific patterns previously reported with the prototype assay, in the same population [17,21]. CRMP-2 + YB-1 and LDH-A + STI1 were identified here as additional combinations associated with transcendence. Clearly, some pairwise combinations are more useful than others in discriminating the affected from control populations and a small panel of five pairwise combinations can identify the large majority of transcendents. While other five-autoantibody combination panels may also have discriminatory value, a table of the top five combinations with the highest sensitivity will thus be used for future cross-validation for development of a final test table for clinical implementation.

The multiplexed ELISA assay comprised of the pairwise protein combinations identified here appears to have potential for clinical implementation in a narrow clinical use case setting to identify a small subset of already high-risk mothers whose biomarker pattern implies very high autism risk, rather than utilizing additional pairwise autoantibody combinations that maximize sensitivity for all cases. That framing explains why the optimization was driven by near-perfect specificity and very high PPV, with detection rates (sensitivity in this context) across all autism cases only of secondary importance as this is a distinct subset of autism with a specific etiology. The key point elucidated here is that the MARA autoantibody assay can identify a substantial group of mothers with very high risk for an autism diagnosis in their children, but the assay does not distinguish mothers at lower risks (<0.9) from TD mothers. This is consistent with the hypothesis that the assay detects a MARA subset of autism.

Thus, this prototype clinical test for MARA presented here is intended to provide information as to whether a child or intended offspring is in fact at a very high likelihood of autism, compared to their already elevated risk based on their early clinical signs, or a mother having had a prior child with autism. Early indication of a clinical diagnosis of autism may allow meaningful earlier interventions which may alter the cascade of presymptomatic interactions and alter the impact of autism.

Early intervention in autistic children may have a significant impact on subsequent development [34–39]. One recent prospective study from Australia provides encouraging evidence for this possibility, as it showed that infants who received preemptive intervention at age three years had lower odds of meeting diagnostic criteria for autism than those who received usual care (7% versus 21%), with the number needed to treat of seven participants [34]. American Academy of Pediatrics guidelines today call for screening for autism and other developmental disabilities at 18 and 24 months of age, and thus a positive MARA test result in those who screen as at risk could predict with a high degree of likelihood an ultimate diagnosis of autism [40]. In that context, a “watch-and-wait” approach may be undesirable for the family and the child.

The EMA study had demonstrated that the autoantibodies were present in a subset of mothers of children with autism during their pregnancy with the affected child [21]. Here we report for the first time on the use of this assay in mothers with a previously diagnosed autistic child who were prospectively followed during a subsequent pregnancy, before the clinical evaluation of their subsequent child (MARBLES). A transcendent population was successfully identified in this context with strong potential predictive value and specificity. Such mothers may also benefit from the clinical application of this assay for pre-conceptual informed decision making. The empiric expectation of 19-20% of autism [41] often results in reproductive stoppage after an affected child [42], and a positive result in this context would suggest a much higher recurrence risk. While families may find the tests’ high predictive value for recurrence a discouraging reason to proceed with subsequent children, this high predictive value can also be viewed by families as an encouraging reason to proceed with subsequent children in that a subsequent child now recognized at very high risk may benefit from very early intervention even in the early pre-symptomatic stage of autism. As has been observed with autosomal recessive disorders, a second potentially affected child is often born before the older sibling is definitively diagnosed, and these children too could benefit from very early intervention when there is evidence of MARA.

Alternatively, gestating in a prenatal environment without the damaging autoantibodies, i.e., through surrogacy, could be an option in the very high risk setting of MARA autoantibodies’ presence. The MARA autoantibody assay is, however, not presently validated for the evaluation of an ongoing pregnancy as the physiological and biochemical changes of pregnancy would need to be carefully evaluated prior to such use.

Pathogenic maternal autoantibodies that cross the placenta during gestation are known to cause congenital heart block in lupus [43], neonatal myasthenia gravis [43,44], and hemolytic disease of the fetus and newborn [45,46]. For these maternal-fetal antibody related disorders, new approaches to remove pathogenic autoantibodies, prevent their placental transfer, or mitigate their effects are becoming increasingly feasible. A test for MARA could potentially be a companion diagnostic for such interventions. One potential intervention is IVIG, which has been widely and safely used, with variable efficacy, in relevant clinical settings, including during pregnancy for primary infertility and recurrent pregnancy loss [47–55].

Monoclonal antibodies targeting the FcRn receptor can prevent the transfer of maternal alloantibodies to the fetus and facilitate rapid elimination of pathogenic autoantibodies [55,56]. Several are already in use for autoimmune disorders, such as myasthenia gravis [44], and a clinical trial for use in pregnancy for hemolytic disease of the fetus and newborn and fetal-neonatal alloimmune thrombocytopenia is now in Phase III [45,46]. Alternative strategies to lower or eliminate maternal autoantibodies by selective targeting and degradation of such autoantibodies are also in development [55–58] One limitation of the current study is that while independently analyzed with a clinically validated assay, the discovery study population is the same as that used in the previous academic publications conducted using the “feasibility” assay, albeit with an expanded subsequent enrollment, which increased the power of the analyses.

While power was not an issue for the CHARGE discovery cohort with 420 unique case subjects, the small number of participants in MARBLES is somewhat limiting but still sufficient for an early external validation set. Following development of a finalized test table and performance validation by cross validation detailed in a sister publication [59], a large external validation study is undergoing recruitment which will allow further evaluation of performance of the MARA autoantibody pairwise combinations.

Specificity and PPV were defined at 100% based on the definition of the thresholds of the transcendent population being both autoantibodies above all control participants’ levels, and claimed performance at this stage represents an extrapolation from this with calculated likely confidence intervals. Prevalence adjustment is necessary for clinically interpretable PPV and NPV and we used here the reported prevalence of autism diagnosis in the two high-risk cohort populations. These two cohorts however closely approximate the intended-use groups for the clinical assay at this time. Future studies will determine performance characteristics directly in populations with other prevalences for broader application.

There is as yet no locked classifier against which to assess the potential clinical value of this assay, however the observations reported here provide the basis to sequentially move to this stage. This model presently identified participants in the data set with the highest level of risk for autism while maximizing the number of positive samples detected and minimizing the number of false positive results. The demonstration of a very high-risk subset of mothers with transcendent pairwise combinations of autoantibodies is a novel discovery pointing to the strong prospect for derivation of a robust final test table, able be deployed clinically. Further analysis is presented in a sister presentation, in which the CHARGE and MARBLES datasets are combined to generate a single test table for both intended uses (IU1 and IU2), consisting of a collection of biomarker pairs, or combinations of these with their associated thresholds [59]. This test table presented in the sister presentation was evaluated using stratified 20-fold cross-validation, with preprocessing performed within each training fold and metrics aggregated across folds to determine the final performance characteristics of a potential clinical test.

The discovery and validation processes reported here highlight the potential for clinical validity of this assay with expected favorable performance characteristics in a relevant portion of autism cases. This in turn may allow a rationale for clinical or future pharmacologic/biologic interventions to mitigate the impact of these maternal autoantibodies.

## Data Availability

All data produced in the present study are available upon reasonable request to the authors

## Author Contributions

Conceptualization, E.R.W.; Methodology, ERW, JvW, M.M.; Validation, M.M.J.S., Formal Analysis, M.M., E.R.W.; Investigation, ERW, M.M.; Resources, J.S., J.VdW., and ERW. Data Curation, M.M. and E.R.W.; Writing – Original Draft Preparation, E.R.W.; Writing – Review & Editing, M.M., J.VdW., E.R.W., M.P.; Project Administration, M.M., E.R.W., M.P.,and J.VdW.; Funding Acquisition, M.M., E.R.W., M.P.,and J.VdW.

## Institutional Review Board Statement

All materials used in this study used existing *de-identified* data/specimens where the investigator did not have access to private identifiable information and no contact with the subjects. Biospecimens used in this study were obtained in accordance with the Declaration of Helsinki, and approved by the Institutional Review Board of the University of California, Davis Protocol ID# 226028, Revised 02/26/2024, and all participants provided informed consent prior to inclusion in the study. Use of biospecimens from the San Diego Blood Bank was waived under WCG IRB Tracking # 20211794 approved 4/22/21.

## Informed Consent Statement

Informed consent for participation was obtained from all subjects involved in the study at the originating institutions, but the use here was under waived approval for de-identified specimens. The study adheres to the scope of research allowed by the original consent.

## Data Availability Statement

The raw data supporting the conclusions of this article will be made available by the authors on request.

## Acknowledgments

The authors would like to thank: MARAbio’s investors including parents of children on the autism spectrum who sought to understand the biological cause of their child’s autism, participants in the UC Davis CHARGE and MARBLES studies, Drs. Mags McInerney and Jessica Barlow for assay development and laboratory services, Stuart Liu-Mayo for review of data methodology, and Dana Barberio of Edge Bioscience Communications for her contribution in editing the manuscript.

## References

1. CDC. Autism Spectrum Disorder. Data and Statistics on Autism Spectrum Disorder. Available online: https://www.cdc.gov/autism/data-research/index.html (accessed on August 20, 2026).

2. Shaw, K.A.; Williams, S.; Patrick, M.E.; Valencia-Prado, M.; Durkin, M.S.; Howerton, E.M.; Ladd-Acosta, C.M.; Pas, E.T.; Bakian, A.V.; Bartholomew, P.;, et al. Prevalence and Early Identification of Autism Spectrum Disorder Among Children Aged 4 and 8 Years -Autism and Developmental Disabilities Monitoring Network, 16 Sites, United States, 2022. MMWR Surveill Summ 2025, 74, 1–22, doi:10.15585/mmwr.ss7402a1.

3. Goines, P.E.; Croen, L.A.; Braunschweig, D.; Yoshida, C.K.; Grether, J.; Hansen, R.; Kharrazi, M.; Ashwood, P.; Van de Water, J. Increased midgestational IFN-gamma, IL-4 and IL-5 in women bearing a child with autism: A case-control study. Mol Autism 2011, 2, 13, doi:10.1186/2040-2392-2-13.

4. Careaga, M.; Van de Water, J.; Ashwood, P. Immune dysfunction in autism: a pathway to treatment. Neurotherapeutics 2010, 7, 283–292, doi:10.1016/j.nurt.2010.05.003.

5. Croen, L.A.; Braunschweig, D.; Haapanen, L.; Yoshida, C.K.; Fireman, B.; Grether, J.K.; Kharrazi, M.; Hansen, R.L.; Ashwood, P.; Van de Water, J. Maternal mid-pregnancy autoantibodies to fetal brain protein: the early markers for autism study. Biol Psychiatry 2008, 64, 583–588, doi:10.1016/j.biopsych.2008.05.006.

6. Braunschweig, D.; Golub, M.S.; Koenig, C.M.; Qi, L.; Pessah, I.N.; Van de Water, J.; Berman, R.F. Maternal autism-associated IgG antibodies delay development and produce anxiety in a mouse gestational transfer model. J Neuroimmunol 2012, 252, 56–65, doi:10.1016/j.jneuroim.2012.08.002.

7. Braunschweig, D.; Ashwood, P.; Krakowiak, P.; Hertz-Picciotto, I.; Hansen, R.; Croen, L.A.; Pessah, I.N.; Van de Water, J. Autism: maternally derived antibodies specific for fetal brain proteins. Neurotoxicology 2008, 29, 226–231, doi:10.1016/j.neuro.2007.10.010.

8. Saunders, N.R.; Liddelow, S.A.; Dziegielewska, K.M. Barrier mechanisms in the developing brain. Front Pharmacol 2012, 3, 46, doi:10.3389/fphar.2012.00046.

9. Saunders, N.R.; Dziegielewska, K.M.; Mollgard, K.; Habgood, M.D. Physiology and molecular biology of barrier mechanisms in the fetal and neonatal brain. J Physiol 2018, 596, 5723–5756, doi:10.1113/JP275376.

10. Hodges, H.; Fealko, C.; Soares, N. Autism spectrum disorder: definition, epidemiology, causes, and clinical evaluation. Transl Pediatr 2020, 9, S55–S65, doi:10.21037/tp.2019.09.09.

11. Camacho, J.; Jones, K.; Miller, E.; Ariza, J.; Noctor, S.; de Water, J.V.; Martinez-Cerdeno, V. Embryonic intraventricular exposure to autism-specific maternal autoantibodies produces alterations in autistic-like stereotypical behaviors in offspring mice. Behav Brain Res 2014, 266, 46–51, doi:10.1016/j.bbr.2014.02.045.

12. Jones, K.L.; Pride, M.C.; Edmiston, E.; Yang, M.; Silverman, J.L.; Crawley, J.N.; Van de Water, J. Autism-specific maternal autoantibodies produce behavioral abnormalities in an endogenous antigen-driven mouse model of autism. Mol Psychiatry 2020, 25, 2994–3009, doi:10.1038/s41380-018-0126-1.

13. Bruce, M.R.; Couch, A.C.M.; Grant, S.; McLellan, J.; Ku, K.; Chang, C.; Bachman, A.; Matson, M.; Berman, R.F.; Maddock, R.J.;, et al. Altered behavior, brain structure, and neurometabolites in a rat model of autism-specific maternal autoantibody exposure. Mol Psychiatry 2023, 28, 2136–2147, doi:10.1038/s41380-023-02020-3.

14. Scattoni, M.L.; Crawley, J.; Ricceri, L. Ultrasonic vocalizations: a tool for behavioural phenotyping of mouse models of neurodevelopmental disorders. Neurosci Biobehav Rev 2009, 33, 508–515, doi:10.1016/j.neubiorev.2008.08.003.

15. Ku, K.M.; Weir, R.K.; Silverman, J.L.; Berman, R.F.; Bauman, M.D. Behavioral Phenotyping of Juvenile Long-Evans and Sprague-Dawley Rats: Implications for Preclinical Models of Autism Spectrum Disorders. PLoS One 2016, 11, e0158150, doi:10.1371/journal.pone.0158150.

16. Kalueff, A.V.; Stewart, A.M.; Song, C.; Berridge, K.C.; Graybiel, A.M.; Fentress, J.C. Neurobiology of rodent self-grooming and its value for translational neuroscience. Nat Rev Neurosci 2016, 17, 45–59, doi:10.1038/nrn.2015.8.

17. Ramirez-Celis, A.; Becker, M.; Nuno, M.; Schauer, J.; Aghaeepour, N.; Van de Water, J. Risk assessment analysis for maternal autoantibody-related autism (MAR-ASD): a subtype of autism. Mol Psychiatry 2021, 26, 1551–1560, doi:10.1038/s41380-020-00998-8.

18. Braunschweig, D.; Krakowiak, P.; Duncanson, P.; Boyce, R.; Hansen, R.L.; Ashwood, P.; Hertz-Picciotto, I.; Pessah, I.N.; Van de Water, J. Autism-specific maternal autoantibodies recognize critical proteins in developing brain. Transl Psychiatry 2013, 3, e277, doi:10.1038/tp.2013.50.

19. Edmiston, E.; Jones, K.L.; Vu, T.; Ashwood, P.; Van de Water, J. Identification of the antigenic epitopes of maternal autoantibodies in autism spectrum disorders. Brain Behav Immun 2018, 69, 399–407, doi:10.1016/j.bbi.2017.12.014.

20. Ramirez-Celis, A.; Edmiston, E.; Schauer, J.; Vu, T.; Van de Water, J. Peptides of neuron specific enolase as potential ASD biomarkers: From discovery to epitope mapping. Brain Behav Immun 2020, 84, 200–208, doi:10.1016/j.bbi.2019.12.002.

21. Ramirez-Celis, A.; Croen, L.A.; Yoshida, C.K.; Alexeeff, S.E.; Schauer, J.; Yolken, R.H.; Ashwood, P.; Van de Water, J. Maternal autoantibody profiles as biomarkers for ASD and ASD with co-occurring intellectual disability. Mol Psychiatry 2022, 27, 3760–3767, doi:10.1038/s41380-022-01633-4.

22. Angkustsiri, K.; Fussell, J.J.; Bennett, A.; Schauer, J.; Ramirez-Celis, A.; Hansen, R.L.; Van de Water, J. Pilot Study of Maternal Autoantibody-Related Autism. J Dev Behav Pediatr 2022, 43, 465–471, doi:10.1097/DBP.0000000000001100.

23. Hertz-Picciotto, I.; Croen, L.A.; Hansen, R.; Jones, C.R.; van de Water, J.; Pessah, I.N. The CHARGE study: an epidemiologic investigation of genetic and environmental factors contributing to autism. Environ Health Perspect 2006, 114, 1119–1125, doi:10.1289/ehp.8483.

24. McLellan, J.; Croen, L.A.; Iosif, A.M.; Ashwood, P.; Yoshida, C.; Berger, K.; Van de Water, J. Differences in mid-gestational and early postnatal neonatal cytokines and chemokines are associated with patterns of maternal autoantibodies in the context of autism. Cereb Cortex 2024, 34, 50–62, doi:10.1093/cercor/bhae082.

25. 25. Van de Water, J.; Wassman, E.R. Chapter 7 - Maternal autoantibodies and fetal neurodevelopment, in Clinical Reproductive Immunology; Karen E. Racicot, G.M., Ed.; Academic Press: 2026.

26. Bagnall-Moreau, C.; Spielman, B.; Brimberg, L. Maternal brain reactive antibodies profile in autism spectrum disorder: an update. Transl Psychiatry 2023, 13, 37, doi:10.1038/s41398-023-02335-3.

27. McInerney, M.; Hurley, B.; Barkow, J.; Menning, K.; Nicolace, J.; Schauer, J.; Van de Water, J.; Wassman, E.R. Analytical Validation of an ELISA Assay for Maternal Autoantibody-Related Autism. Diagnostics (Basel) 2026, 16, doi:10.3390/diagnostics16111665.

28. Hertz-Picciotto, I.; Schmidt, R.J.; Walker, C.K.; Bennett, D.H.; Oliver, M.; Shedd-Wise, K.M.; LaSalle, J.M.; Giulivi, C.; Puschner, B.; Thomas, J.;, et al. A Prospective Study of Environmental Exposures and Early Biomarkers in Autism Spectrum Disorder: Design, Protocols, and Preliminary Data from the MARBLES Study. Environ Health Perspect 2018, 126, 117004, doi:10.1289/EHP535.

29. 29. Snow, A. Social Communication Questionnaire, in Encyclopedia of Autism Spectrum Disorders, Volkmar, F.R. (editor); Springer, New York, NY: 2013.

30. Croen, L.A.; Grether, J.K.; Hoogstrate, J.; Selvin, S. The changing prevalence of autism in California. J Autism Dev Disord 2002, 32, 207–215, doi:10.1023/a:1015453830880.

31. Ojala, M.; Garriga, G.C. Permutation Tests for Studying Classifier Performance. Journal of Machine Learning Research 2010, 11, 1833–1863.

32. Clopper, C.J.; Pearson, E.S. The use of confidence or fiducial limits illustrated in the case of the binomial. Biometrika 1934, 26, 404–413, doi:10.1093/biomet/26.4.404.

33. Mercaldo, N.D.; Lau, K.F.; Zhou, X.H. Confidence intervals for predictive values with an emphasis to case-control studies. Stat Med 2007, 26, 2170–2183, doi:10.1002/sim.2677.

34. Whitehouse, A.J.O.; Varcin, K.J.; Pillar, S.; Billingham, W.; Alvares, G.A.; Barbaro, J.; Bent, C.A.; Blenkley, D.; Boutrus, M.; Chee, A.;, et al. Effect of Preemptive Intervention on Developmental Outcomes Among Infants Showing Early Signs of Autism: A Randomized Clinical Trial of Outcomes to Diagnosis. JAMA Pediatr 2021, 175, e213298, doi:10.1001/jamapediatrics.2021.3298.

35. Szatmari, P.; Chawarska, K.; Dawson, G.; Georgiades, S.; Landa, R.; Lord, C.; Messinger, D.S.; Thurm, A.; Halladay, A. Prospective Longitudinal Studies of Infant Siblings of Children With Autism: Lessons Learned and Future Directions. J Am Acad Child Adolesc Psychiatry 2016, 55, 179–187, doi:10.1016/j.jaac.2015.12.014.

36. MacDuffie, K.E.; Estes, A.M.; Peay, H.L.; Pruett, J.R., Jr.; Wilfond, B.S. The Ethics of Predicting Autism Spectrum Disorder in Infancy. J Am Acad Child Adolesc Psychiatry 2021, 60, 942–945, doi:10.1016/j.jaac.2021.01.006.

37. Frazier, T.W.; Klingemier, E.W.; Anderson, C.J.; Gengoux, G.W.; Youngstrom, E.A.; Hardan, A.Y. A Longitudinal Study of Language Trajectories and Treatment Outcomes of Early Intensive Behavioral Intervention for Autism. J Autism Dev Disord 2021, 51, 4534–4550, doi:10.1007/s10803-021-04900-5.

38. Brefort, E.; Saint-Georges-Chaumet, Y.; Cohen, D.; Saint-Georges, C. Two-year follow-up of 90 children with autism spectrum disorder receiving intensive developmental play therapy (3i method). BMC Pediatr 2022, 22, 373, doi:10.1186/s12887-022-03431-x.

39. Williamson, I.O.; Elison, J.T.; Wolff, J.J.; Runge, C.F. Cost-Effectiveness of MRI-Based Identification of Presymptomatic Autism in a High-Risk Population. Front Psychiatry 2020, 11, 60, doi:10.3389/fpsyt.2020.00060.

40. Lipkin, P.H.; Macias, M.M.; Council On Children With Disabilities, S.O.D.; Behavioral, P. Promoting Optimal Development: Identifying Infants and Young Children With Developmental Disorders Through Developmental Surveillance and Screening. Pediatrics 2020, 145, doi:10.1542/peds.2019-3449.

41. Bernie, C.; Williams, K.; Graham, F.; May, T. Coaching While Waiting for Autism Spectrum Disorder Assessment: Protocol of a Pilot Feasibility Study for a Randomized Controlled Trial on Occupational Performance Coaching and Service Navigation Support. JMIR Res Protoc 2021, 10, e20011, doi:10.2196/20011.

42. Ozonoff, S.; Young, G.S.; Carter, A.; Messinger, D.; Yirmiya, N.; Zwaigenbaum, L.; Bryson, S.; Carver, L.J.; Constantino, J.N.; Dobkins, K.;, et al. Recurrence risk for autism spectrum disorders: a Baby Siblings Research Consortium study. Pediatrics 2011, 128, e488–495, doi:10.1542/peds.2010-2825.

43. Madhusudan, D.; Raju, A.; Vijaya, N. Correlation of Maternal Autoantibodies with Fetal Congenital Heart Block. J Obstet Gynaecol India 2016, 66, 112–116, doi:10.1007/s13224-015-0813-7.

44. Lindroos, J.L.V.; Bjork, M.H.; Gilhus, N.E. Transient Neonatal Myasthenia Gravis as a Common Complication of a Rare Disease: A Systematic Review. J Clin Med 2024, 13, 1136, doi:10.3390/jcm13041136.

45. Zhu, L.N.; Hou, H.M.; Wang, S.; Zhang, S.; Wang, G.G.; Guo, Z.Y.; Wu, J. FcRn inhibitors: a novel option for the treatment of myasthenia gravis. Neural Regen Res 2023, 18, 1637–1644, doi:10.4103/1673-5374.363824.

46. Moise, K.J., Jr.; Ling, L.E.; Oepkes, D.; Tiblad, E.; Verweij, E.; Lopriore, E.; Smoleniec, J.; Sachs, U.J.; Bein, G.; Kilby, M.D.;, et al. Nipocalimab in Early-Onset Severe Hemolytic Disease of the Fetus and Newborn. N Engl J Med 2024, 391, 526–537, doi:10.1056/NEJMoa2314466.

47. Maisonneuve, E.; Panchaud, A.; Baud, D. Inhibiting IgG in Hemolytic Disease of the Fetus. N Engl J Med 2024, 391, 563–567, doi:10.1056/NEJMe2401020.

48. 48. Arumugham, V.B.; Rayi, A. NIH. National Library of Medicine. StatPearls. Intravenous Immunoglobulin (IVIG). Available online: https://www.ncbi.nlm.nih.gov/books/NBK554446/ (accessed on March 2, 2026).

49. Peero, E.K.; Banjar, S.; Khoudja, R.; Ton-Leclerc, S.; Beauchamp, C.; Benoit, J.; Beltempo, M.; Dahan, M.H.; Gold, P.; Kadoch, I.J.;, et al. Intravenous immunoglobulin for patients with unexplained recurrent implantation failure: a 6-year single center retrospective review of clinical outcomes. Sci Rep 2024, 14, 3876, doi:10.1038/s41598-024-54423-z.

50. Yamada, H.; Deguchi, M.; Saito, S.; Takeshita, T.; Mitsui, M.; Saito, T.; Nagamatsu, T.; Takakuwa, K.; Nakatsuka, M.; Yoneda, S.;, et al. Intravenous immunoglobulin treatment in women with four or more recurrent pregnancy losses: A double-blind, randomised, placebo-controlled trial. EClinicalMedicine 2022, 50, 101527, doi:10.1016/j.eclinm.2022.101527.

51. Mustafa, H.J.; Sambatur, E.V.; Pagani, G.; D’Antonio, F.; Maisonneuve, E.; Maurice, P.; Zwiers, C.; Verweij, J.; Flood, A.; Shamshirsaz, A.A.;, et al. Intravenous immunoglobulin for the treatment of severe maternal alloimmunization: individual patient data meta-analysis. Am J Obstet Gynecol 2024, 231, 417–429 e421, doi:10.1016/j.ajog.2024.03.044.

52. Connery, K.; Tippett, M.; Delhey, L.M.; Rose, S.; Slattery, J.C.; Kahler, S.G.; Hahn, J.; Kruger, U.; Cunningham, M.W.; Shimasaki, C.;, et al. Intravenous immunoglobulin for the treatment of autoimmune encephalopathy in children with autism. Transl Psychiatry 2018, 8, 148, doi:10.1038/s41398-018-0214-7.

53. Nimmerjahn, F.; Ravetch, J.V. Anti-inflammatory actions of intravenous immunoglobulin. Annu Rev Immunol 2008, 26, 513–533, doi:10.1146/annurev.immunol.26.021607.090232.

54. Tjon, A.S.; van Gent, R.; Geijtenbeek, T.B.; Kwekkeboom, J. Differences in Anti-Inflammatory Actions of Intravenous Immunoglobulin between Mice and Men: More than Meets the Eye. Front Immunol 2015, 6, 197, doi:10.3389/fimmu.2015.00197.

55. Sultan, Y.; Kazatchkine, M.D.; Maisonneuve, P.; Nydegger, U.E. Anti-idiotypic suppression of autoantibodies to factor VIII (antihaemophilic factor) by high-dose intravenous gammaglobulin. Lancet 1984, 2, 765–768, doi:10.1016/s0140-6736(84)90701-3.

56. Nagelkerke, S.Q.; Kuijpers, T.W. Immunomodulation by IVIg and the Role of Fc-Gamma Receptors: Classic Mechanisms of Action after all? Front Immunol 2014, 5, 674, doi:10.3389/fimmu.2014.00674.

57. Peter, H.H.; Ochs, H.D.; Cunningham-Rundles, C.; Vinh, D.C.; Kiessling, P.; Greve, B.; Jolles, S. Targeting FcRn for immunomodulation: Benefits, risks, and practical considerations. J Allergy Clin Immunol 2020, 146, 479–491 e475, doi:10.1016/j.jaci.2020.07.016.

58. Devanaboyina, S.C.; Khare, P.; Challa, D.K.; Ober, R.J.; Ward, E.S. Engineered clearing agents for the selective depletion of antigen-specific antibodies. Nat Commun 2017, 8, 15314, doi:10.1038/ncomms15314.

59. Wassman, E.R.; Martin, M.; McInerney, M. Cross Validation Assessment of Clinical Performance Characteristics of a Test Table for the Risk of Maternal Autoantibody-Related Autism in High-Risk Populations (manuscript in preparation; to be submitted). 2026.

